# Electroacupuncture on Modifying Inflammatory Levels of Cytokines and Metabolites in Stroke Patients

**DOI:** 10.1101/2023.04.11.23288440

**Authors:** Arriagada Rios Sandra, Liao Yi Fang, Yu Chen Lee, Ming-Kuei Lu, Sheng-Ta Tsai, Ben-Arie Eyal, Wen-Chao Ho

## Abstract

**Introduction:** The use of electroacupuncture (EA) in post-ischemic stroke and rehabilitation has been the subject of numerous studies; however, the effect of EA on cholesterol metabolites has not been thoroughly investigated. The inflammatory response in stroke has been associated with serum cholesterol, low HDL-Cc, and high LDL-Cc levels, and early intervention has been linked to improved post-stroke rehabilitation. This study aimed to assess the impact of EA on early ischemic stroke as a modulator of total cholesterol, HDL-c, and LDL-c in the blood, its anti-inflammatory effect, and its effect on pain and stroke scales in patients in the first few days after the onset of stroke.

**Data Access Statement:** The datasets generated during the current study are available from the corresponding author on reasonable request

**Material and Method:** A total of 90 patients with acute ischemic stroke and a first-time diagnosis of stroke will be randomized into one of three groups: an EA group, a sham EA group, and a sensory control group. All patients will receive the interventions three times a week for a total of six sessions over two weeks. Outcome measurements will include blood tests for total cholesterol, triglycerides, HDL with HDL-c cholesterol, LDL and LDL-c cholesterol, along with Visual Analog Scale (VAS), National Institutes of Health Stroke Scale (NIHSS), and Barthel Index (BI).

**Expected Outcome:** This study will help determine the effect of EA on ischemic stroke recovery, focusing on metabolic changes in patients with early stage stroke. EA treatment might modify risk indices (HDL-c), maintain or control (LDL-c), and generate localized reperfusion of the vascular areas involved in stroke.

**Discussion:** This randomized controlled trial will determine the ability of EA to support early stroke ischemic injury and neuro-endothelium damage, which could lead to a faster stroke recovery in stroke scales, and reveal whether the mechanism of EA is associated with a reduced inflammatory process via modulation of the levels of total cholesterol, HDL-c, LDL-c, and triglycerides. The results of this study will be of significant value in the treatment of ischemic stroke and could lead to more effective and personalized stroke rehabilitation therapies.

**Trial registry:** registered study protocol on www.clinicaltrial.gov (NCT05734976)

## Introduction

Ischemic stroke is characterized by multiple interconnected neuropathophysiological cascades, including intense inflammatory processes (1). Treatment for these patients requires early intervention, which should be started as soon as 4– 5 h after the infarction, as this has been associated with better clinical outcomes in ischemic stroke, which is strongly related to timely revascularization (2). There is sufficient evidence that post-ischemic inflammation is associated with acute disruption of the blood-brain barrier (BBB), vasogenic edema, and hemorrhagic transformation (3). The inflammatory response in stroke can be associated with high serum cholesterol, high LDL-c, and low HDL-c levels (4).

Endothelial injury is the first step in the pathogenesis of vascular lesions, in which lipid plaques and fibrous elements accumulate inside arteries. Endothelial lesions triggers inflammation with increased adhesiveness and activation of leukocytes and platelets, accompanied by the production of cytokines, chemokines, vasoactive molecules, and growth factors (5,6). If the serum level of oxidized low-density lipoprotein (Ox-LDL) is increased, macrophage uptake is also increased in early vascular plaque lesions and may be mediated by interleukin-6, which increases foam cell formation in macrophages, which in turn increases the contraction of blood vessels and induces hypertrophy and hyperplasia of blood vessels and smooth muscle cells (VSMC) (7,8).

In contrast, HDL-c exerts the opposite effects as a direct endothelial protector, through the production of nitric oxide molecules, which have antioxidant, anti-inflammatory, and antithrombotic effects. By raising its plasma levels, HDL-c can stimulate endothelial repair processes, which involve mobilization and promotion of the endothelial repair capacity of endothelial progenitor cells, stimulating antiatherosclerotic effects. The therapeutic properties of HDL play a crucial role in the regulation of vascular tone and vasodilation, which acts directly on the endothelium (9,10).

The pattern of blood stagnation syndrome in traditional Chinese medicine theory generates blood flow disturbance, vascular dysfunction, and endothelial damage, a process similar to that occurring in stroke (11). One technique used in Chinese medicine is electroacupuncture (EA) stimulation, which is commonly used in the clinical treatment of ischemic stroke (12).

The protective effects of EA against endothelial dysfunction have been verified. These properties include the modulation of the autonomic nervous system (ANS), endothelial nitric oxide synthase (eNOS), and NOS. The modulation of the ANS by suppressing the sympathetic nervous system is especially important in the treatment of cardiovascular diseases. Li et al. also showed that acupuncture needling at ST36 (the point used in our study) activated NO synthesis, which is the main modulator of local blood flow and vascular tone. EA reduces the size of the cerebral infarct by increasing eNOS activity, which increases local perfusion (13).

The effectiveness of EA is closely related to effective intervention of risk factors. For example, animal model experiments have confirmed that EA has therapeutic effects on hypertension, hypotension, myocardial ischemia, and several types of arrhythmias (14,15). EA can activate the sympathetic inhibitory system in the brain, resulting in the release of endogenous opioids, γ-amino-n-butyric acid, and serotonin. These mediators can inhibit sympathetic neurons in the nucleus paragigantocellularis lateralis (PGL) of the rostral ventrolateral medulla oblongata (rVLM), which is responsible for maintaining blood pressure (BP) and cardiovascular integration reflexes. (16,17)

This study aimed to assess the ability of electroacupuncture (EA) to support patients with early ischemic stroke, which could lead to faster recovery from stroke, with respect to total cholesterol, HDL-c, and LDL-c levels in the blood and their anti-inflammatory effects in patients with stroke. This study also aimed to evaluate the impact of AEs as modifiers of analgesia and stroke scales. Measurement of post-AD serum levels (total cholesterol, HDL-c, LDL-c, and triglycerides) will provide adequate information on the anti-inflammatory role of EA on these risk factors, blood vessels, and the vascular endothelium. We hypothesized that EA could modulate the metabolic response to acute inflammation by modulating total cholesterol, HDL-c, and LDL-c levels in patients with ischemic stroke.

## METHOD

### Design and Setting

This single-blind randomized controlled trial was conducted between September 2022 and September 2023 in the Department of Neurology at China Medical University Hospital, Taichung, Taiwan. Our trial was approved by the institutional review board (IRB) of China Medical University Hospital (CMUH111-REC1-132), and the study protocol was registered at www.clinicaltrial.gov (NCT05734976).

#### Objectives

This study aimed to determine the therapeutic efficacy of EA in promoting the recovery of patients with early ischemic stroke. We will further explore the mechanism of EA associated with the serum levels of the main metabolites derived from cholesterol, which increase the level of HDL-c and decrease the level of LDL-c, affecting the cellular anti-inflammatory response.

#### Participants

Ninety patients diagnosed with ischemic stroke (ICD10 I63-I66) and admitted to the hospital for the first time will be included in this randomized clinical trial. Eligible subjects will be required to provide informed consent and will be randomly assigned to one of the three groups: EA, sham EA, and sensory control. All patients will receive interventions three times a week for a total of six sessions over two weeks. To reduce potential bias in this study, both the investigator and patients will be blinded to the group allocation. **See Figure 1**.

**Figure 1.**
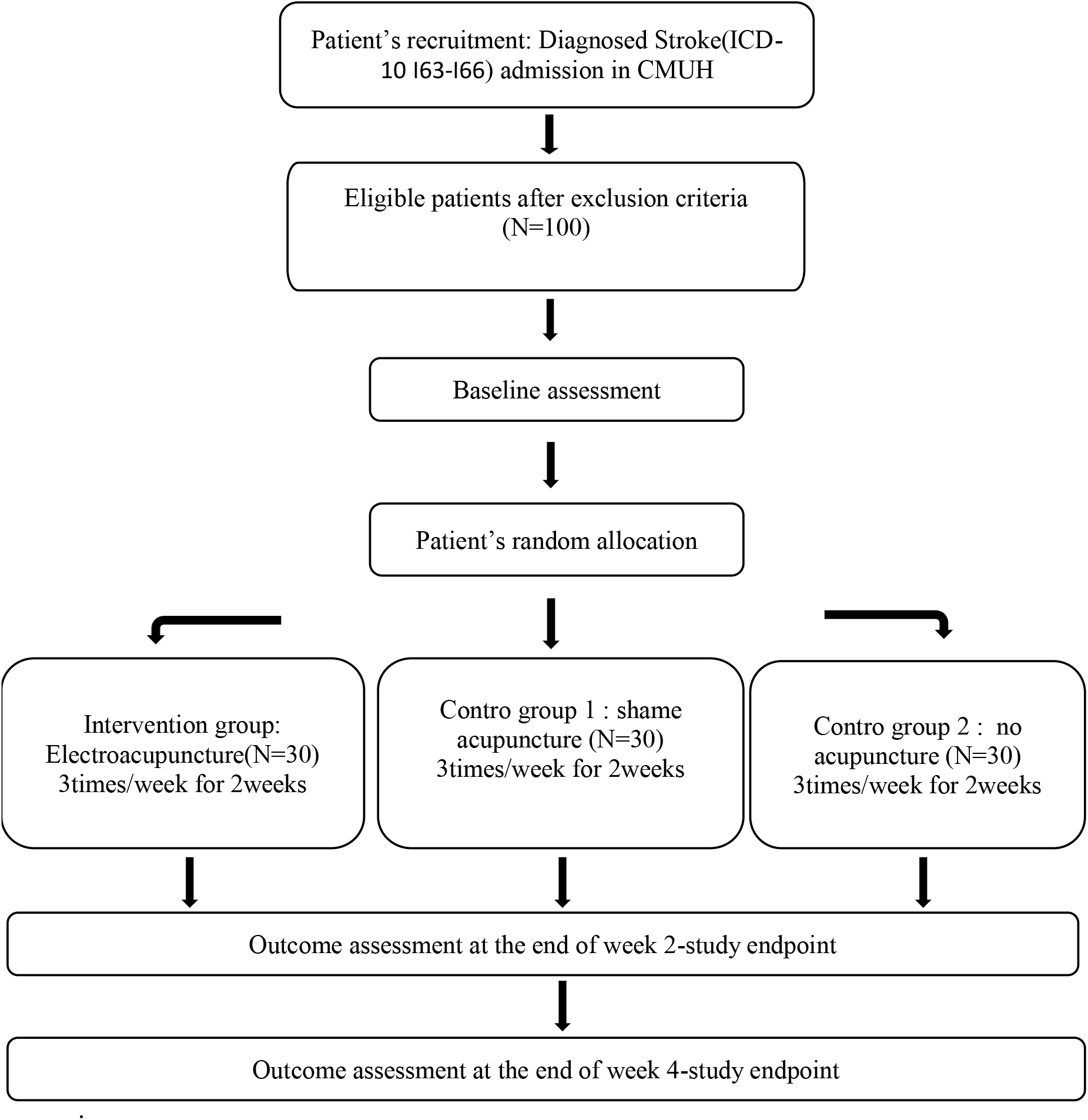
Study flow chart. begins with the recruitment of the patient: diagnosed stroke (ICD-10 I63-I66) admission to CMUH. Continuing with eligible patients after the exclusion criteria, patients were randomly assigned into three intervention groups: (a) AE group, (b.) sham EA, (and c): sensory control group. Finally, the evaluation of the results at the end of week 4 was the: endpoint of the study.

#### Participants

Ninety patients diagnosed with first-time ischemic stroke who meet our inclusion criteria were included in this single-blind, randomized clinical trial, conducted at the Department of Neurology, China Medical University Hospital, Taichung, Taiwan. Patients will be included immediately after stroke, and the intervention will begin as soon as possible.

### Inclusion criteria

1. Diagnosis of acute ischemic stroke.
2. First-time stroke diagnosis.
3. Age between 40 and 80 years old.
4. Both genders.

### Exclusion criteria

1. Diagnosis of malignant cancer.
2. Traumatic event.
3. Patients undergoing immunosuppressive therapy.
4. History of drug abuse.
5. Pregnancy.
6. Patients with a cardiac pacemaker.

### Recruitment Strategies

The research team, consisting of neurologists and acupuncture specialists, met once a month to evaluate the selection and recruitment of patients, as well as to monitor the admission of patients who met the inclusion criteria through direct conversation with their families.

### Informed consent

Before the patient’s legal guardian signed the informed consent form, the study director explained the patients’ rights, objectives, characteristics, and risks. Patients could withdraw from the study at any time after providing their informed consent. In this case, the patient’s data will be retained for the final analysis, unless the patient requests otherwise. Informed consent was obtained in the traditional Chinese language for easy reading by the patients and/or their families. Number IC: CF-V4,20230208.

### Randomization and allocation concealment

Ninety patients will be randomized in a 1:1:1 ratio into three groups: EA, EA sham, and control. Randomization was performed using the IBM SPSS Statistics version 22 software (SPSS Inc., Chicago, IL, USA). A simple computer-based randomization method, without stratification, was implemented. Randomization will occur prior to patient enrollment, and the file and randomization information will be accessible only to the administrator. Ninety non-transparent sealed envelopes were prepared, with patient assignments for each group. At the time of patient inclusion, the doctor will choose an envelope at random and write the patient’s name and date of birth, after which the acupuncturist will apply the interventions in the envelope. This process ensured that both the investigator and patients were blinded to the group assignment. In some acupuncture studies, positive results are thought to be due to a placebo effect, often related to a sensory reaction. In this study, we controlled for the placebo effect through two control groups: a sham EA group and a no-EA, non-penetrating sensory control group. (Figure 2)

**Figure 2.**
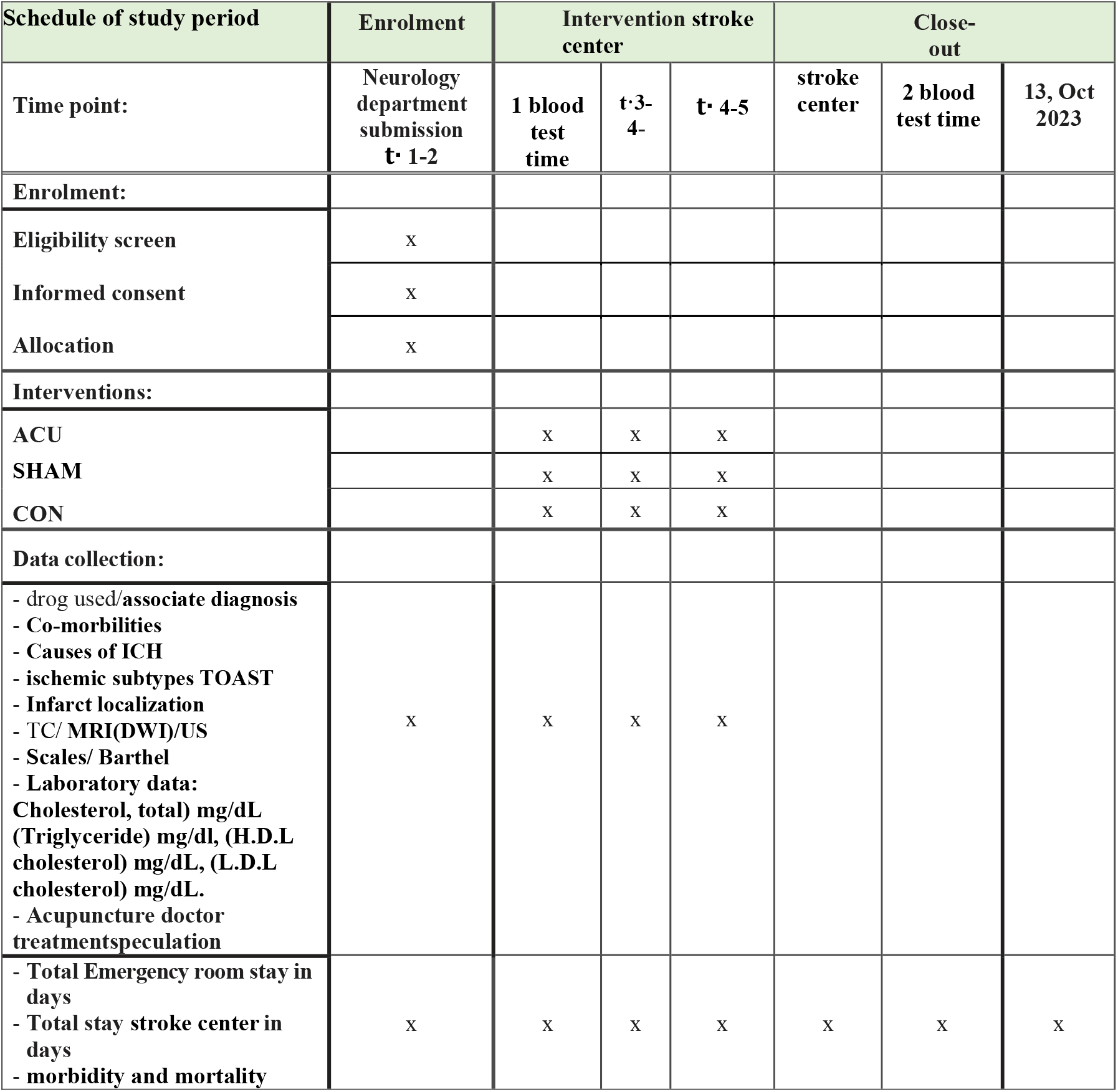
Test schedule of study period. enrollment schedule, eligibility assessment, informed consent, allocation, interventions. Data collection: (x) Main outcome measurement, (t·) time point. (50).

### Interventions

#### EA Group

This group will receive treatment at 8 acupoints including Baihui (GV-20), Sishencong (EX-HN1), Quchi (LI11), Hegu (LI4), Neiguan (PC6), Waiguan (TE5), Yanglingquan (GB34), and Zusanli (ST36) during each EA session using a 2 Hz frequency. This group will receive EA applied to selected acupuncture points on both arms and legs, as well as on the scalp. All procedures will be performed with disposable needles (CLOUD AND DRAGON Medical Device Co. Ltd.) that are 0.25 mm in diameter (32 gauge) and 44 mm in length. Needle insertion in this group will follow the TCM acupuncture style and will penetrate the muscular level of the body acupoints and the subcutaneous level of the scalp acupoints. The needles were rotated in both directions to reach the sensation of “De Chi” before turning on the Ching Ming EA machine (Model 05 B ISO900113415). The EA frequency used was proximal distal, and the scalp dose was 2 Hz. (photo 1)

#### Sham EA Group

This group will receive treatment at sham points located 1 cm lateral to six acupoints: Quchi (LI11), Hegu (LI4), Neiguan (PC6), Waiguan (TE5), Yanglingquan (GB34), and Zusanli (ST36). The sham points are not known acupuncture points and are not located on the acupuncture meridians, making them sham acupoints. All needles will be inserted to a depth of 0.5 cm with disposable needles without needle rotation and without “De Chi.” The needles were then connected to and EA machine at a frequency of 2 Hz. The needle type and EA machine used were identical to those used in the EA group.

#### Sensory Control Group

This group will receive tactile touch stimuli at six acupuncture points: Quchi (LI11), Hegu (LI4), Neiguan (PC6), Waiguan (TE5), Yanglingquan (GB34), and Zusanli (ST36), without needle penetration and, without the use of regular needles or EA. This group served as the sensory control group. The needle plastic tube (without a needle) was lightly pressed onto the skin for a few seconds at a few points. This might give patients the impression that they are receiving acupuncture, and might serve to measure the placebo effect. (All information on the acupuncture points is shown in Table 1.)

**Table 1.** Acupuncture points used in this study. The name, number, location, dose and a scheme are observed according STandards for Reporting Interventions in Clinical Trials of Acupuncture (STRICTA): extending the CONSORT Statement (33,34)

| Acupoint name | Acupoint number | Location<br>C5-T11 |
| --- | --- | --- |
| Baihui 百會 | (GV-20)<br>DM 20<br>D 2 Hertz | At the junction of a line connecting the apices of the ears and the midline<br>N; frontal, branches of the trigeminal A external carotid branches |
| Sishencong 四神聰 | (EX-HN1)<br>D 2 Hertz<br>bilateral | A group of four points, each located 1 cun from → Du-20 (anterior, posterior and lateral)<br>N; frontal, branches of the trigeminal A external carotid branches |
| Quchi 曲池 | (LI11)<br>D 2 Hertz<br>bilateral | With the elbow flexed, on the lateral end of the elbow crease.<br>N radial<br>A recurrent radial |
| Hegu 合谷 | (LI4)<br>D 2 Hertz<br>bilateral | On the radial aspect of the hand, between the 1st and 2nd metacarpal bones<br>N median<br>A radial |

D: electroacupuncture dosage, N: nerve, A: artery
| Acupoint name | Acupoint number | Location<br>L3-L4 |
| --- | --- | --- |
| Neiguan 內關 | (PC6)<br>D 2 Hertz<br>bilateral | 2 cun proximal to the anterior wrist joint space<br>N median<br>A radial |
| Yanglingquan 陽陵泉 | (GB34)<br>D 2 Hertz<br>bilateral | In the depression anterior and inferior to the head of the fibula<br>N fibular common<br>A tibial anterior |
| Zusanli<br>足三里 | (ST36)<br>D 2 Hertz<br>bilateral | lateral to the anterior crest of the tibia, on the tibialis anterior muscle.<br>N nervio recurrente articular, fibular<br>A circunfleja fibular |
| Waiguan<br>外關 | (TE5)<br>SJ 5<br>D 2 Hertz<br>bilateral | 2 cun proximal to the dorsal wrist joint, between the radius and the ulna<br>N median, superficial branches<br>A radial superficial |

### Drug Treatment

Different treatment protocols were used for these patients, depending on the degree of complexity. These include intravenous (IV) thrombolysis, mechanical thrombectomy with intra-arterial thrombolysis (IA), and reperfusion windows for patients with acute ischemic stroke. The reperfusion window is the time window for reperfusion therapies to begin within the first 0 5 h. Intravenous (IV) thrombolysis with tissue plasminogen activator (tPA, alteplase) is the standard of care in the treatment of acute ischemic stroke in current clinical practice, with the time window extended up to 4.5 hours after the onset of symptoms. During this study, western medical treatment protocols will continue as usual, and doctors will proceed with routine and post-stroke care. The study interventions will also be considered. At the onset of the study interventions, the patients were hemodynamically stable.

### Outcome measures

The main outcome measurements were laboratory data from the blood samples, including total cholesterol (mg/dL), triglycerides (mg/dL), HDL cholesterol (mg/dL), and LDL cholesterol (mg/dL). This study aimed to evaluate the modulation of serum cholesterol levels, especially HDL-c and LDL-c levels, which are known to cause acute inflammation in patients with stroke. These measurements were performed twice: before and after EA treatment

### Secondary Outcomes

Neurological assessments will be used with patients, including the visual analog scale (VAS) for general pain measures, National Institutes of Health Stroke Scale (NIHSS), and Barthel Index (BI). These evaluations will be conducted at the beginning, middle (two weeks), and end of the treatment.

### Timing of each measurement

The first blood sample will be obtained when the patient is admitted to the CMUH Neurology Service in the morning, and the second comparable sample will be obtained after the patient has finished treatment (approximately one month later).

### Adverse events

During the informed consent discussion, the patient’s legal guardian will be informed of the possible side effects of acupuncture, including minor bleeding, allergic reactions to alcohol, bruising, fainting, local inflammation, and skin irritation. Any serious adverse events that occur during the trial will be immediately reported to the principal investigator and the ethics committee of China Medical University Hospital. If a serious adverse event occurs, the patient group will be unassigned, the patient will receive appropriate treatment, and the ethics committee will decide whether the study will continue. Details of the adverse events, including the number of events in a patient, the outcome of the patient’s recovery, the location of adverse events, and the selected acupuncture point, will be recorded.

### Sample size calculation

Sample size estimation for longitudinal studies will be performed using G*Power 3.1.9.7, a statistical program designed to estimate statistical power and effect size. This study considered two samples before and after EA treatment, evaluating fixed effects, special main effects, and interactions. Sensitivity, effect size, and power were assessed. The power F-test G multiple linear regression was used, with a fixed model, R2 deviation from zero, complete α=0.05, power of verification=0.8, and a total of five independent variables. The calculated sample size was 74 patients, which resulted in a power of 0.87.

### Data management

Data will be collected from the online medical records of the CMUH, as well as from a manual case report sheet for each patient. Access to the collected data from medical records will be secure, and only authorized study personnel will be permitted to use it for research purposes. The manual case report sheets will be kept in a secure location in the department office and will be destroyed five years after the trial completion. The primary investigator of the study will ensure that the patients meet the study criteria and that the data are correctly collected. Any changes to the study protocol will require approval from the CMUH IRB. Personal information about potential and enrolled participants will be collected and maintained to protect confidentiality, and patient names or identifying information will not be used. Only the digital data were used in this study.

### Statistical analysis

Statistical analyses were performed using IBM SPSS Statistics (version 22.0; (SPSS Inc., Chicago, IL, USA). Data are presented as percentages (n%) for categorical data and mean ± standard deviation (SD) for continuous data. Patients will be randomly assigned to the EA, Sham EA, and sensory control groups. To generate statistical operational information, a generalized estimating equation (GEE) approach was used. GEE considers the within-subject correlation of data (longitudinal studies of data before and after EA treatment) and models the average response. When applicable, a one-way analysis of variance (ANOVA) was performed. The objective was to make inferences about the population by considering intrasubject correlations. The comparison of the three groups (estimate for longitudinal studies: quantitative variable) of blood levels of serum cholesterol and metabolites before and after the study interventions in patients with GEE stroke will indicate the average response.

## Discussion

This study aimed to investigate the influence of EA on total cholesterol, HDL-c, LDL-c, triglycerides, and BMI, allowing for an accurate comparison of patients’ inflammatory processes that cause stroke events and hinders their recovery. EA can induce localized reperfusion of vascular areas involved in stroke (18, 19). By working with metabolic rates (blood analysis), the results can be compared with Western medicine equipment, enabling the observation and evidence of the benefits of acupuncture.

A decrease in blood fluidity can cause disorders in the circulatory system, such as arterial sclerosis or embolization, causing damage to vascular endothelial cells due to hypertension, chronic inflammation, decreased flexibility of blood vessels due to hyperlipidemia and aging, and weak blood cells. The antihypertensive effect of EA is associated with oxidative stress and IGF-I in animal models of RF-induced hypertension (20, 21).

EA provides favorable conditions for the reconstruction of neurovascular units in the early stages of a brain attack. EA increases angiogenesis by increasing endothelial growth factor (VEGF) and, improving collateral circulation. Angiogenesis is initiated rapidly to maintain the integrity of the blood-brain barrier and ensure energy supply to neurons (22, 23).

Ischemia/reperfusion injury leads to microglial activation through circulatory inflammation, exacerbating brain damage. EA can interfere with this process via the p38 mitogen-activated protein kinase (MAPK) and ERK pathways and downregulates the expression of IL-1β, IL-6, and TNF-α, thereby reducing the inflammatory response. One example of the anti-inflammatory actions of acupuncture on ST36 and the associated changes in mediators of inflammation has clinical benefits in alleviating inflammation through several mechanisms, including activation of the vagus nerve, receptor 4 (TLR4) signaling/NF-κB, macrophage polarization, activation of the mitogen-activated protein kinase (MAPK) pathway, and the cholinergic anti-inflammatory pathway (24, 25, 26).

The findings of this study will help determine the effects of EA and metabolic changes ion patients diagnosed with early -stage stroke. Additionally, the design of this study will contribute to the analysis of blood test results, allowing for an accurate comparison of individual HDL-C, LDL-C, and triglyceride levels and ratios, thus improving our understanding of the neurovascular origin of this phenomenon (27,28). The repair of the neuro-endothelium, determined by the increase in vascular flow, ensures adequate pain management, especially during the rehabilitation stages (29).

When there is an exchange of information about clinical procedures, such as in this study involving metabolic changes in associated inflammatory states, specific planning and enrollment times are required. Our follow-up was performed only in the Taiwanese population, at a single specialized center for ischemic stroke. In the future, it will be necessary to associate studies on cholesterol metabolites with specific inflammatory markers, such as cytokines, PCR, or VHS, for clinical and therapeutic projections (30,31,32).

## Data Availability

Data Access Statement: The datasets generated during the current study are available from the corresponding author on reasonable request

## Author contribution

**Analysis and interpretation of the data**: Ho Wen-Chao and Eyal Ben Arie,

**Conceptualization**: Sandra Arriagada R., Lee Yu Chen, Ming-Kuei Lu

**Data curation**: Eyal Ben-Arie.

**Design**: Ming-Kuei Lu, Lee Yu Chen, Sandra Arriagada R.

**Formal analysis**: Ho Wen-Chao.

**Funding acquisition**: Lee Yu Chen.

**Investigation:** Sandra Arriagada R.

**Methodology:** Lee Yu Chen, Sandra Arriagada R., Eyal Ben-Arie

**Patients enrollment**: Ming-Kuei Lu, Sheng-Ta Tsai, Liao Yi Fang

**Project administration:** Ming-Kuei Lu, Lee Yu Chen.

**Resources:** Lee Yu Chen.

**Software:** Eyal Ben-Arie, Ho Wen-Chao.

**Supervision:** Lee Yu Chen.

**Trial sponsor**: Sponsor: China Medical University Hospital. Responsible Party: Principal Investigator: Yu-Chen Lee [d5167] Official Title: Chief and Attending Physician, Department of Acupuncture, China Medical University Hospital, Taichung, Taiwan. Affiliations: China Medical University Hospital

**Validation:** Lee Yu Chen, Eyal Ben-Arie

**Visualization:** Sandra Arriagada R.

**Writing – original draft**: Sandra Arriagada R, Eyal Ben-Arie

**Writing – review and editing**: Sandra Arriagada R. Eyal Ben-Arie

**Protocol registration IRB approval number**: CMUH111-REC1-132

**Registered study protocol on:** www.clinicaltrial.gov: NCT05734976

## Acknowledgments

The authors thank the Chinese Medicine Research Center, and the Center for Stroke Patients Medical and China Medical University for supporting this study.

## ABBREVIATIONS

CHOL-t: Total cholesterol level
HDL-c: high-density lipoproteins level
LDL-c: Low-density lipoprotein level
BMI: Body Mass Index
Ox-LDL: Macrophage uptake of oxidized low-density lipoproteins
EA: Electroacupuncture
CMUH: China Medical University Hospital
NO: Nitric oxide (NO) is a signaling molecule that plays a key role in the pathogenesis of inflammation. NO is considered a pro-inflammatory mediator that induces inflammation by overproduction in abnormal situations.
NADPH: NADPH is the reduced form of NADP+; used in anabolic reactions, such as the synthesis of lipids and nucleic acids.
MPO/HOCl: system plays an important role in the elimination of microbes by neutrophils. Furthermore, MPO has been shown to be a local mediator of tissue damage and resulting inflammation in various inflammatory diseases.
LOOH: the accumulation of lipid hydroperoxides
VSMC: vascular smooth muscle cell
IL-6: interleukin-6 is implicated in a large number of acute and chronic diseases associated with inflammation.
ANS: autonomic nervous system
eNOS: endothelial nitric oxide synthase enzyme
IRB: Institutional Review Board

## photo: example of electro acupuncture points

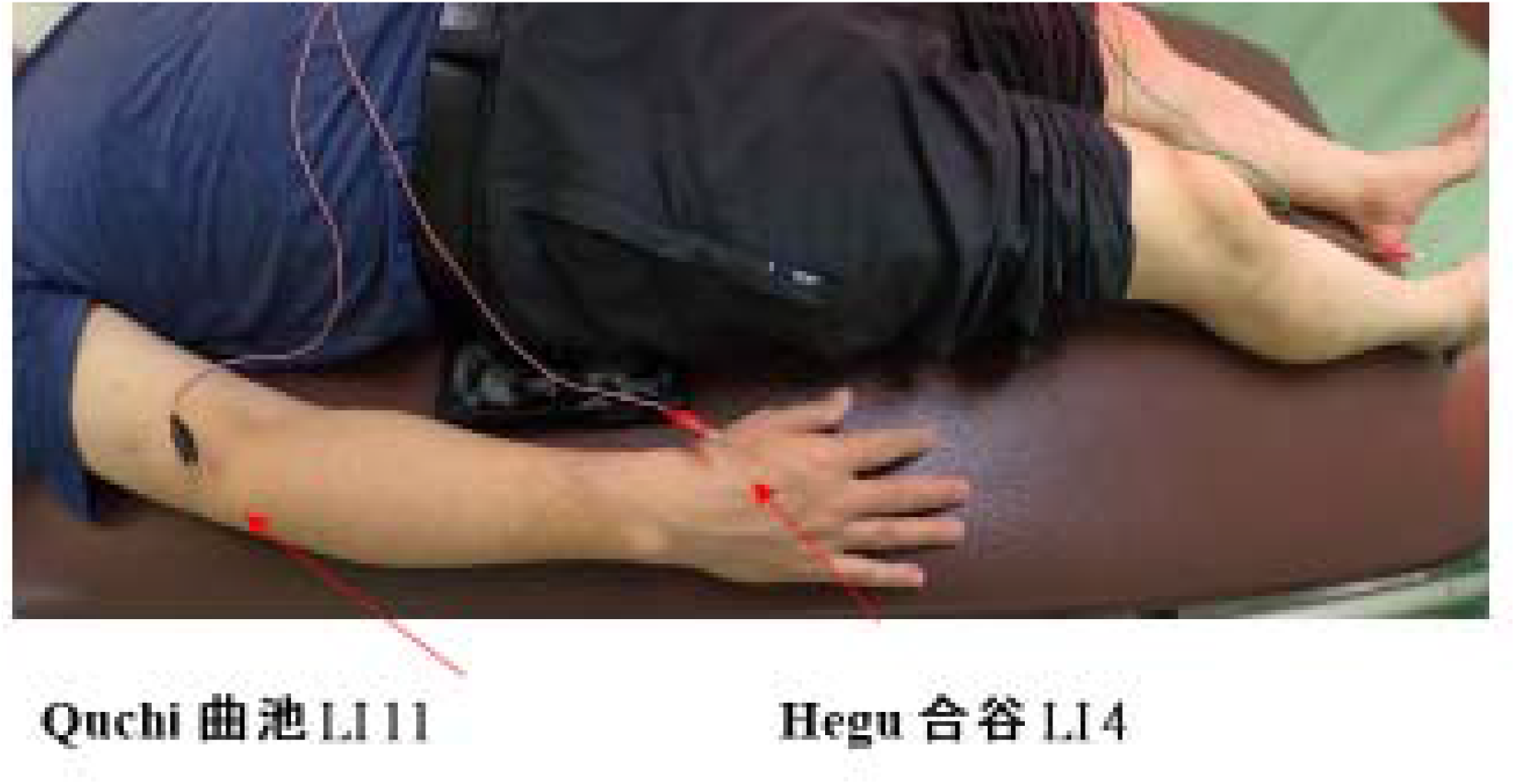

